# Scrub typhus in northern Thailand: a seroprevalence study among Thai and highland populations

**DOI:** 10.64898/2026.08.03.26359547

**Authors:** Carlo Perrone, Meiwen Zhang, Elizabeth M. Batty, Stuart D. Blacksell, Nicholas P. J. Day, Nidanuch Tasak, Ratchadaporn Papwijitsil, Nattida Toonin, Jantana Wongsantichon, Suphasuta Khongpraphan, Yoel Lubell, Thomas J. Peto, Sue J. Lee

## Abstract

**Background:** Scrub typhus is a major cause of febrile illness in Southeast Asia, with particularly high burdens reported in northern Thailand. Environmental exposures and use of preventive measures remain incompletely understood.

**Methods:** We conducted a population-based cross-sectional serosurvey in Chiang Rai province, Thailand in 2023. Adults aged ≥15 years from 74 villages were selected by two-stage random sampling and interviewed regarding demographic characteristics, agricultural and domestic exposures, and preventive behaviours. IgG antibodies against *Orientia tsutsugamushi* were measured using a Luminex xMAP Intelliflex® platform. Mixed-effects logistic regression models stratified by ethnicity (highland people vs Thai) were used to identify factors associated with seropositivity, with village as a random intercept.

**Results:** Among 1,194 participants with valid serology, seroprevalence was higher among highland people than Thai ones (60% vs 13%, p<0.001). Seropositivity increased with age in both groups (p<0.001, and p=0.006), more markedly among highland people. Exposure to forest/high hill (adjusted odds ratio [aOR] 2.87, 95%CI 1.07–7.70) and agroforestry plantations (aOR 1.74, 95%CI 1.04–2.99) were associated with seropositivity among Thai participants, but not among highland people. High-risk occupations were associated with increased odds of seropositivity across both groups, p=0.006 and p=0.043. No clear associations were observed between seropositivity and duration of exposure, domestic/peridomestic risk factors, individual preventive practices, or overall prevention quality. Significant village-level heterogeneity persisted after adjustment, particularly among highland people.

**Conclusions:** Scrub typhus exposure is common in rural Chiang Rai and disproportionately affects highland people; whose risk was poorly explained by classical environmental risk factors, suggesting distinct and incompletely understood exposure pathways. Current prevention strategies based on conventional risk messaging may therefore have limited effectiveness in these high-risk communities.

## Introduction

Scrub typhus, a vector-borne disease, is one of the leading infectious causes of morbidity in rural Southeast Asia (1–3). In the province of Chiang Rai, northern Thailand, 22.5% of acute undifferentiated febrile illness episodes were due to scrub typhus, with farmers from ethnic groups collectively known as highland people (or “hill tribes”, a term considered discriminatory by some) being at higher risk (4, 5).

Trombiculid mite larvae (also known as chiggers) are the vector, and the causative agent is *Orientia tsutsugamushi*, an obligate intracellular bacterium (6, 7). Small vertebrates such as rodents or shrews are typical maintenance hosts, while humans are incidental dead-end hosts (8). Environmental exposure has long been recognised as the main risk factor for scrub typhus infection and risk environments have been broadly categorised into three groups (9): overgrown or neglected fields, fringe environments, and grassy riverbanks. However, cases have been linked to a wide range of habitats, including urban and peri-domestic ones (8, 10, 11); accordingly, not only farmers, forest workers and military personnel, but also hikers and housewives, have been identified as at risk (9, 12–15). In Thailand and other countries, forests have often been associated with increased risk of scrub typhus (16–18).

Human behaviour can influence both the development of environments favourable to chigger-host ecotypes and the risk of being bitten by the vector. Studies have investigated the protective effect of preventive behaviours (e.g. wearing protective clothing, changing and washing regularly, avoiding contact with the naked soil) on scrub typhus (16, 17, 19) but findings have been inconsistent, possibly due to variations in local epidemiology, ecology, and practices (13, 20, 21). Recently, a large study from southern India indicated the possibility of differing risk patterns in high- and low-risk populations (11).

Highland people ethnic groups such as the Akha, Lahu, Karen, Lisu, Shan, Hmong, and Yao; are at increased risk of scrub typhus. To understand their risk profile, a study on environmental and behavioural risk was carried out in Chiang Rai province(20, 22). However, the study was conducted in a single area without any ethnic Thais, limiting its generalizability. We therefore carried out a population-based survey of participants from five districts of Chiang Rai province, without restrictions on ethnicity or occupation, to estimate the effects of exposure to agricultural environments and preventive behaviour on scrub typhus risk.

## Methods

### Design and data collection

Data was collected between October 3^rd^ 2022 and June 22^nd^ 2023. This study was part of the South and Southeast Asia Community-based Trials Network (SEACTN) household survey (HHS) project, whose primary aim was to estimate the prevalence of communicable and non-communicable diseases and their risk factors in Thailand, Bangladesh, and Cambodia. This sub-study was conducted at the Thai site in Chiang Rai province only.

Methods are described in detail elsewhere (23). Briefly, this was a cross-sectional household survey carried out in villages referring to primary care units (PCUs) within the SEACTN (24). The PCUs were purposively selected based on distance from referral hospitals (the farther ones were preferred) and patient volume (those with higher visit volumes were preferred). The sampling frame covered 186 villages, with a total population of 126,315. Participants were selected through two-stage cluster randomised sampling. In the first stage, 75 villages were randomly selected with a likelihood proportional to their population size and in the second stage, five to seven households per village were selected by simple random sampling. All members of the households were invited to participate. For this analysis, participants aged 15 years or older with valid blood samples and laboratory results were included.

### Procedures

Consenting participants from the sampled households were interviewed with a standardised questionnaire, adapted from DHS and WHO STEPS tools and included into an electronic case report form (eCRF) (23). Household heads provided information on ethnicity and household characteristics, including household wealth, which was evaluated by an asset-based questionnaire (EquityTool)(25). Individual participants provided information on their demographic and socioeconomic status (age, sex, education, occupation) and a series of questions on scrub typhus risk during the previous 12 months, including the frequency of A) domestic/ peridomestic risk activities; B) agricultural environment exposure; including rice fields, forest or high hill, agroforestry crops (coffee, tea, or fruit plantations), and dry fields; and C) preventive or risk behaviour (asked only to those who answered they had been exposed to at least one of the agricultural environments listed in B). These questions were chosen based on previously published literature and local, experience-based understanding of scrub typhus risk (16, 20, 26). The variables were analysed dichotomously (present or not present, regardless of frequency) and semi-quantitatively based on frequency.

To examine the overall effect of prevention in relation to other factors and to increase statistical power, the questions in (C) were recoded into a single prevention score, which was then divided into tertiles for analysis. The complete list of questions on scrub typhus is included in Appendix A, and derivation of exposure duration and of the prevention score are presented in Appendix B. The full questionnaire is presented elsewhere (23).

Occupations were categorised into high- (agriculture, day labourer, unemployed or retired) and low-risk (all other occupations. Agricultural environments were also categorized into high-risk (forest/high hill and agroforestry environments) and low-risk (all other environments). High risk occupational and environmental categories were chosen based on local knowledge and literature review (18, 20, 27, 28).

### Serological testing

Antibodies to *O. tsutsugamushi* (among other febrile illness, including rickettsial and non-rickettsial, pathogens) were measured using the Luminex xMAP Intelliflex® (Luminex Corporation, Austin, U.S.A.) platform, which included recombinant 56kDa type-specific antigens of the Karp, Kato, and Gilliam strains. IgG values were reported as the median fluorescent intensity with background subtracted (MFI-bg). Positivity was defined as IgG values to any of the antigens higher than three standard deviations above the mean of the first component of a Gaussian distribution, obtained by fitting the log_10_ transformed MFI-bg to finite mixture models (29).

### Statistical analysis

Statistical analysis was carried out using STATA version 17.0. Variables were summarised using frequencies with percentages or median with interquartile range (IQR), as appropriate. The chi-squared test was used to compare demographic characteristics between highland people and Thai participants and seropositivity across categories of independent variables.

Logistic regression models were used to quantify the association between *O. tsutsugamushi* seropositivity and: (1) exposure to, and duration of exposure in, agricultural environments; (2) exposure to household risk activities; and (3) individual preventive practices and the overall quality of prevention (20, 30). Inclusion of variables in multivariable regression models were guided by a directed acyclic graph (DAG) and reflected the relationship between exposure to risk environments and scrub typhus seropositivity, following general and local epidemiological knowledge (20, 31, 32). The minimum recommended adjustment included occupation, age, and village (Figure 1). To account for the non-linear relationship with seropositivity, age was divided into 15-year catefories. To account for sampling seasonality and differences in geographical, ecological, climatic, and agricultural features (all of which can impact scrub typhus risk), villages were added as a random intercept in a mixed effects model. The median odds ratio (MOR) for the clustering variable (village) was calculated to provide a more easily interpretable estimate of between-cluster variation (33).

**Figure 1.**
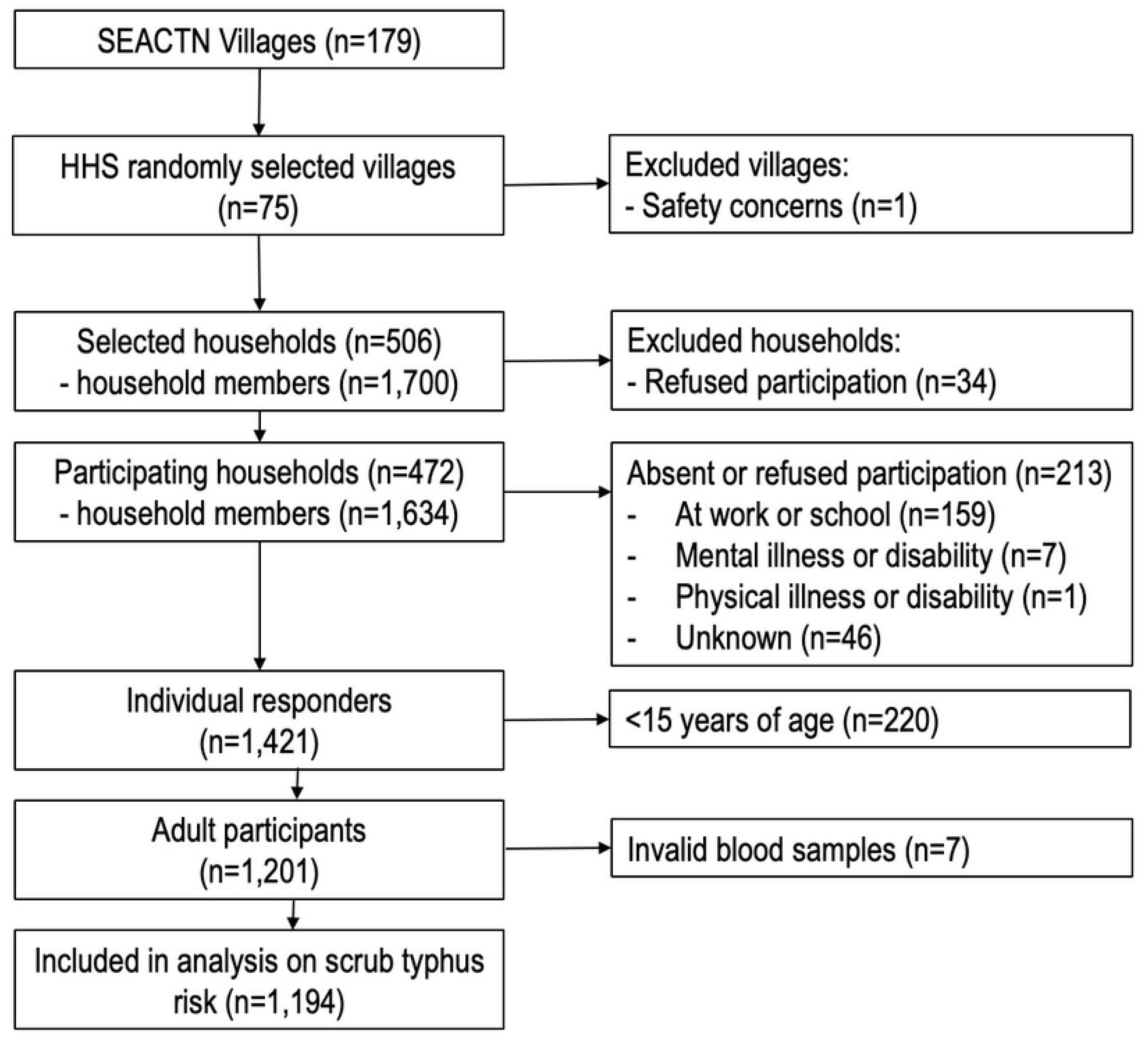
Flowchart of participants in the scrub typhus risk factor analysis of the South and Southeast Asia Community Trials Network (SEACTN) Household Survey.

## Results

Within the 74 participating villages, 506 households were selected and 1201 eligible adults (>=15 years) agreed to participate. Seven participants did not have a valid blood sample; therefore, 1194 were included in this scrub typhus study (**Error! Reference source not found.**). The main reason for nonparticipation, when given, was being at work or at school (n=159, 75%).

**Figure 2.**
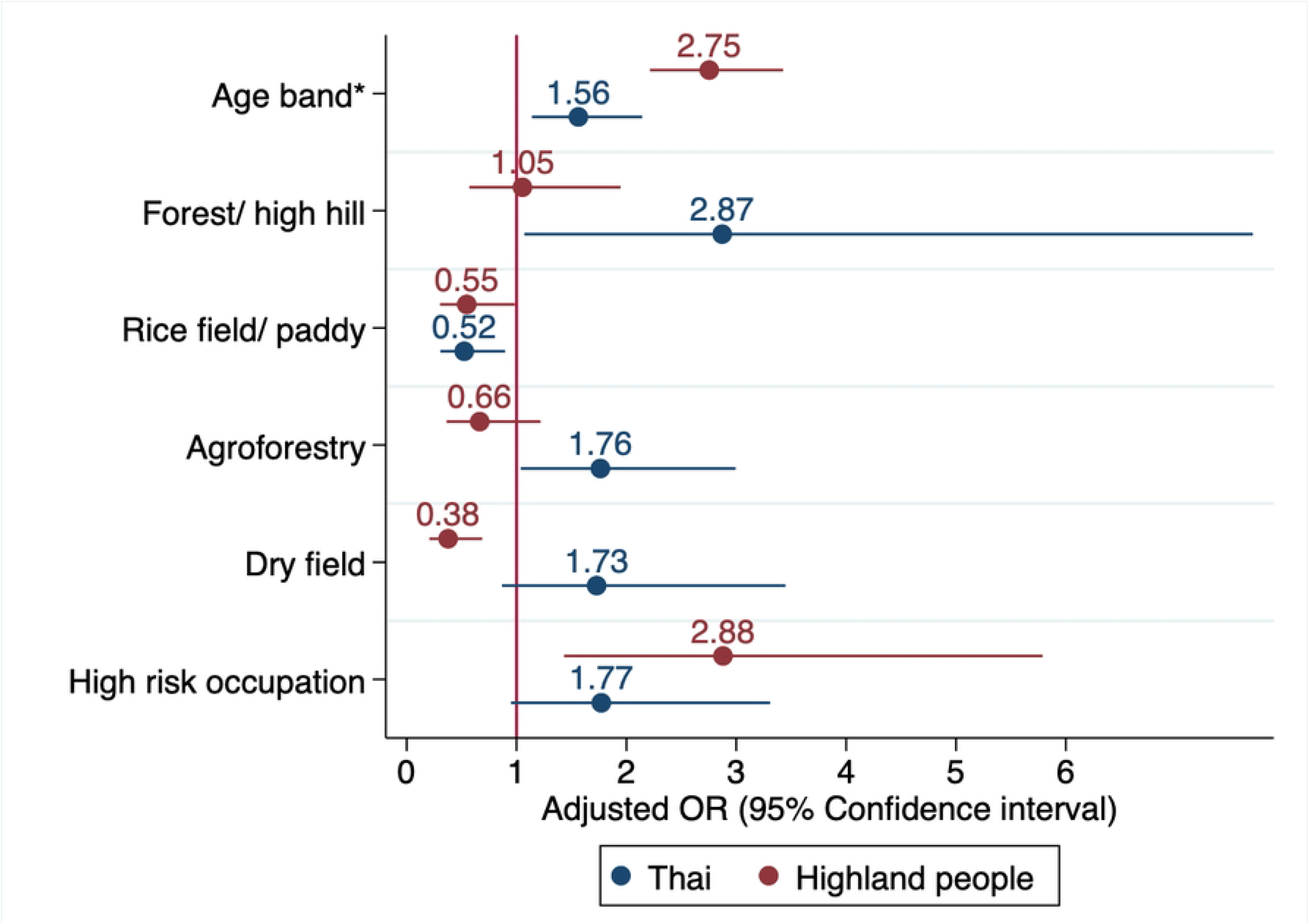
Forest plot from the mixed effects multivariable logistic regression models for highland people (n=481, 291 seropositive) and Thai (N=711, 91 seropositive) participants. The analysis included high-risk occupations, agricultural exposures, and age-bands, using the village as a random intercept. Colours are used to distinguish findings by ethnicity, with blue indicating Thai and red indicating highland people. The dots indicate the adjusted odds ratios, and the bars indicate the 95% confidence intervals. The red vertical line indicates unity (OR=1). *Age bands: ≥15-30, 31-45, 46-60, ≥61, included as a continuous variable. High-risk occupation: Agriculture, day labourer, unemployed or retired

Median age was 55 years (IQR: 43-64). There were fewer younger adults (15-45 years: n=368, 31%) than older adults, and fewer men (n=483, 40%), Table 1. Overall, 456 (38%) participants had never attended school or not completed primary school, and 73.1% (n=870) had high-risk occupations, with 61% (n=730) working in agriculture. More than half of the participants were Thai (n=712, 60%). Ethnicity was clustered by village, with 89% (n=66) of the selected villages consisting only of Thai or only of highland people. Compared to Thai, highland people were younger (median 44 years vs 59), had lower wealth (49% below the two lowest quintiles vs 21%), lower education levels (48% had never attended school vs 7%), and were more often agricultural workers (68% vs 57%).

**Table 1.**
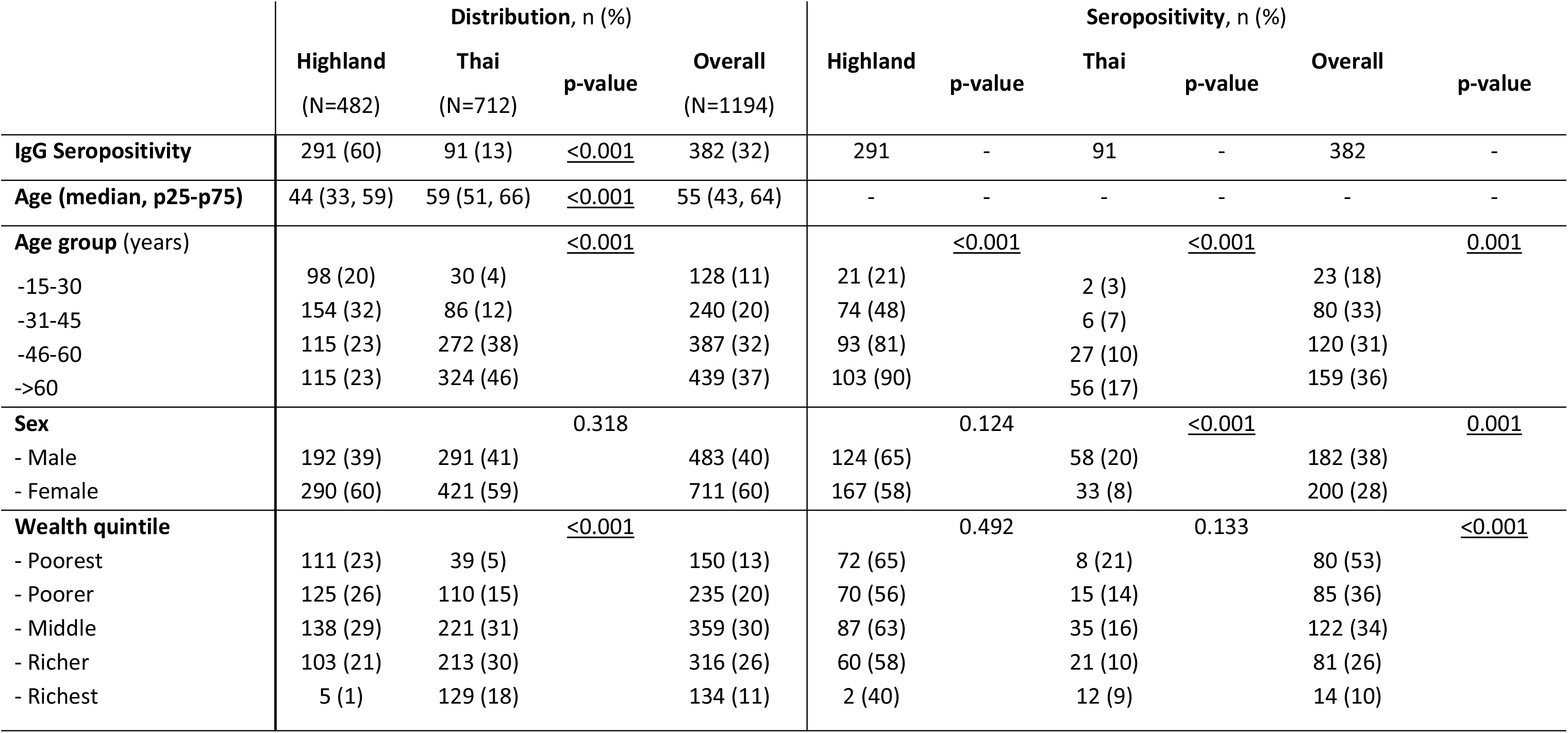
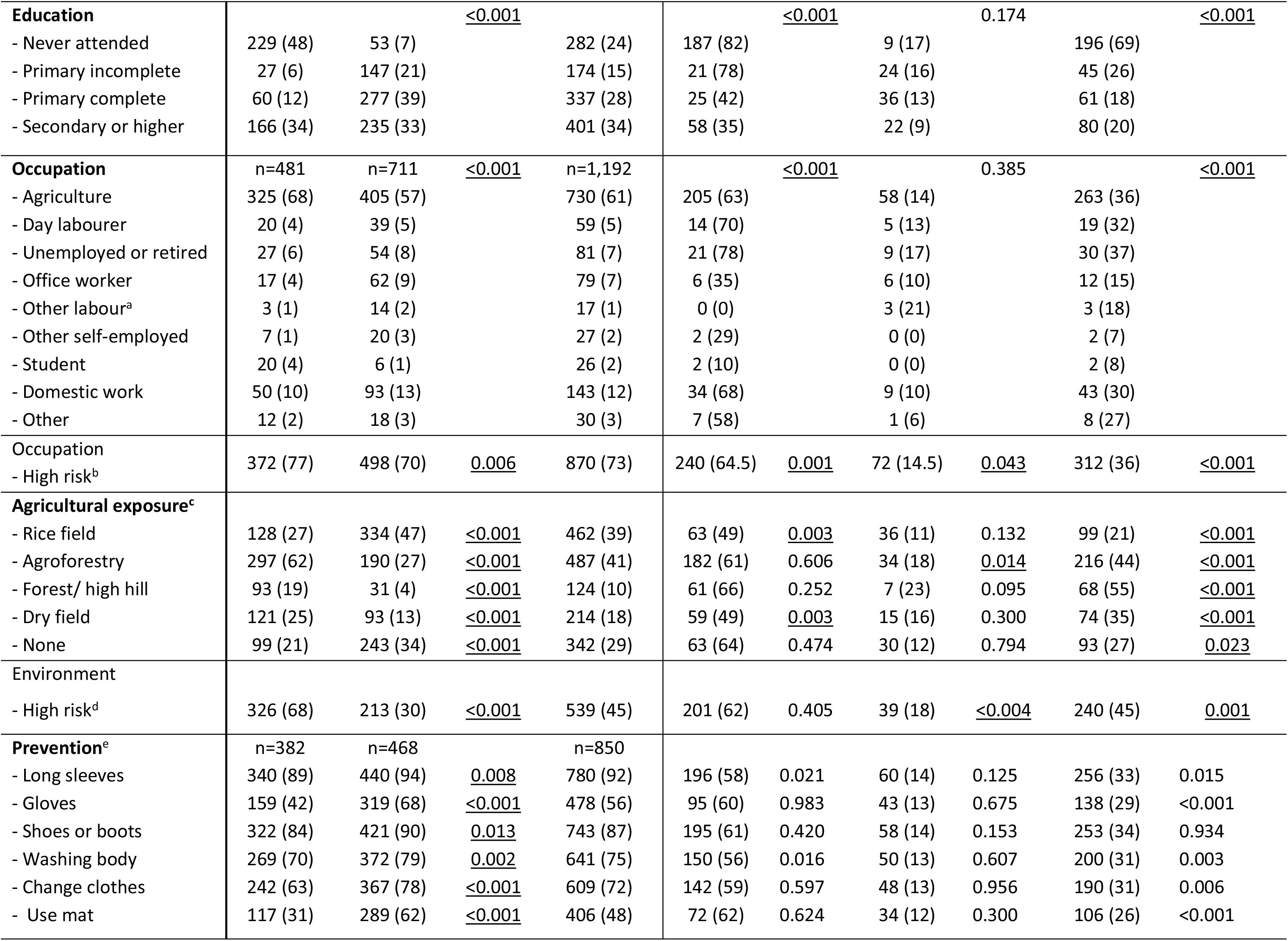
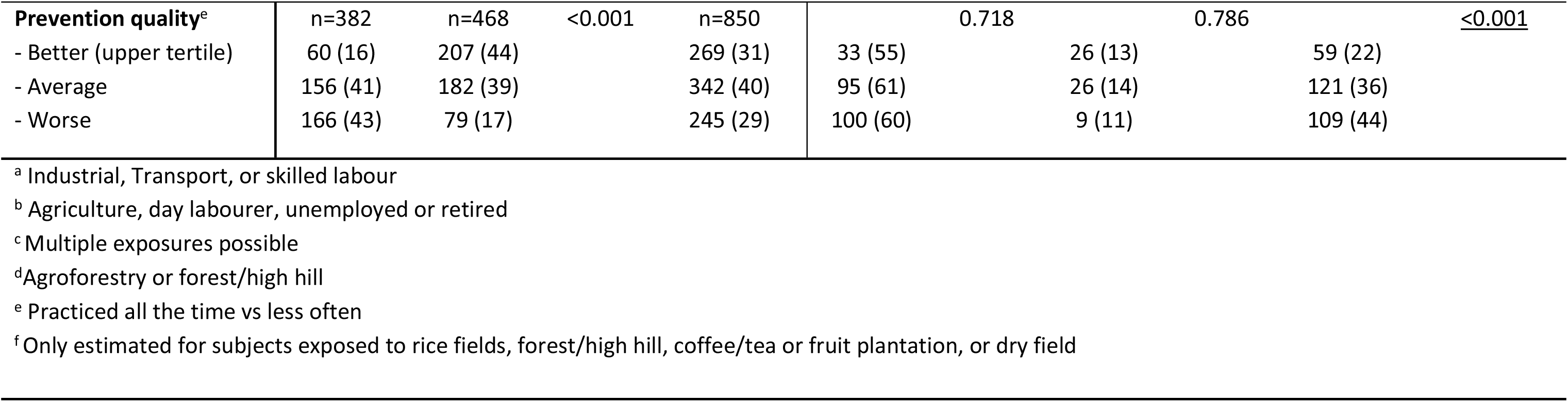
Demographic characteristics, exposure variables, and seropositivity by ethnic group. Numbers are n (%) unless otherwise indicated, in columns for the distribution and in rows for seroprevalence. Distribution is compared between Hill tribe and Thai participants, while seropositivity is compared across variable categories, separately for highland-people, Thai, and all participants. All comparisons were made using the Chi-square test.

The exposure across agricultural environments often overlapped, with 44% of individuals having been exposed to more than one environment (supplementary material, Figure S1). A higher proportion of highland people were exposed to key agricultural environments (79% vs 66%), particularly to agroforestry plantations (62% vs 27%), forest/high hills (19% vs 4%), and dry fields (25% vs 13%); the Thai were more often exposed to rice fields (47% vs 27%).

Questions about prevention were asked only to adults who had visited agricultural environments (n=850) (Table1). Protective measures were well practiced: almost all participants wore long-sleeved shirts (92%, n=780) and shoes or boots (87%, n=743) all the time. However, only 56% (n=478) and 48% (n=406), reported always using gloves or a mat when resting on the naked soil, respectively. Compared with Thai participants, highland people consistently reported lower levels of personal protection and were more likely to fall into the worst prevention-quality tertile (43% vs 17%, p<0.001).

Overall seroprevalence was 32% (n=382) and was higher among highland people (n=291, 60%) than among Thai participants (n=91, 13%). Seroprevalence was also higher among males and those in the lower wealth quintiles or with lower formal education, and lower among office workers (Table 1).

The demographic and exposure differences presented above and in Table 1 caused significant confounding between ethnicity and seropositivity, making a single model unstable; separate models were therefore run for hill-tribe and Thai participants. Observations with missing values were excluded.

Age was significantly associated with increased odds of seropositivity among Thai (aOR 1.56, 95% CI: 1.14-2.14) and highland people (aOR 2.75, 95% CI: 2.21-3.43; Figure 3), per 15-year age increment. Exposure to agricultural environments carried different levels of risk for Thai and highland people. Among both groups, consistent directions for aORs were observed for rice paddy (highland people: 0.55 [95% CI: 0.30, 0.99]; Thai: 0.52 [0.31-0.90]), forest/high hill (highland people: 1.05 [95% CI: 0.57, 1.95]; Thai: 2.87 [1.07-7.70]), and high-risk occupation (highland people: 2.88 [1,43, 5.79]; Thai: 1.77 [0.95, 3.31]), although not all statistically significant. In contrast, the two groups showed opposite trends for agroforestry plantations (highland people: 0.66 [0.36, 1.22]; Thai: 1.76 [1.04, 2.99]) and dry fields (highland people: 0.38 [0.21, 0.69]; Thai: 1.73 [0.87, 3.45]).

**Figure 3:**
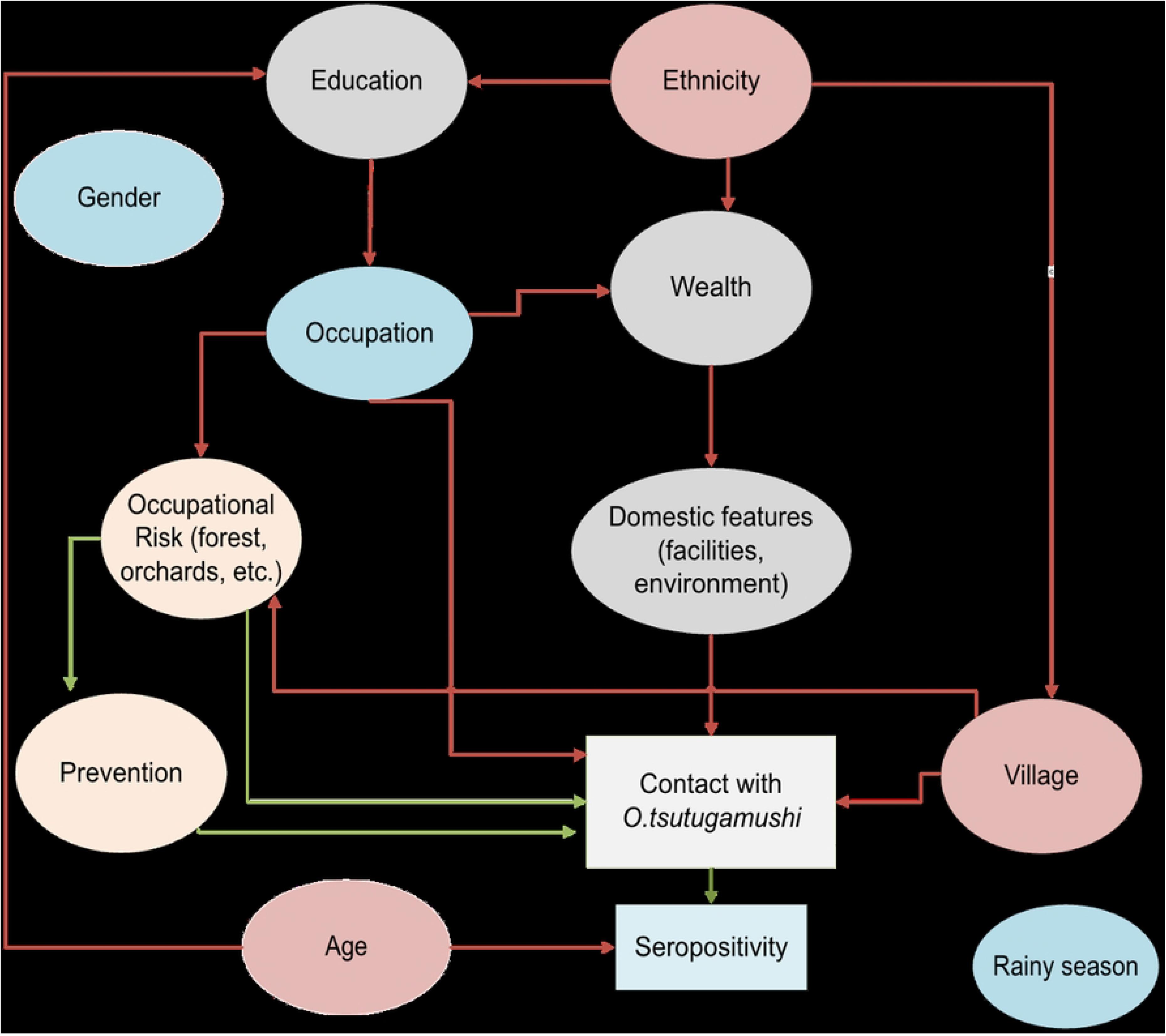
Directed acyclic graph outlining the causal pathways between variables and confounding pathways. Seropositivity is the outcome of interest. Variables in red or purple are confounders, and in orange are the exposures of interest for the analysis. Variables in purple are the ones adjusted for in the mixed effects model. The village was included as a random intercept to account for clustering. Red arrows indicate confounding pathways.

There was no significant, clear, or consistent association between seropositivity and self-reported duration of exposure to agricultural environments in any category, among either highland people or Thai participants (Supplementary Table 1).

None of the preventive measures were associated with seropositivity, nor was the overall quality of prevention, neither among Thai nor among highland people (Supplementary Table 2). Similarly, none of the analysed domestic/peridomestic risk factors (solid fuel use, sitting on the naked flood, infrequent mowing of the house lawn) were associated with seropositivity (Supplementary Table 3). Highland people domestic workers did, however, have high seroprevalence, comparable to participants with high-risk occupations.

Significant village-level random effects indicated substantial heterogeneity in seropositivity across villages, independent of individual-level risk factors. This was more pronounced among highland people, who had a median odds ratio (MOR) of 2.18 (95% CI 1.64, 3.47) in high-versus-low-risk villages, whereas among the Thai, the MOR was 1.87 (95% CI 1.37, 3.34). These findings confirm that residing in a high-risk area remains one of the strongest predictors of risk.

## Discussion

In northern Thailand, research on risk factors for infectious diseases has typically focused on either highland-people-only or Thai–only populations; to the best of our knowledge, this is the first study to include members of both living in the same area. Highland people had a much higher seroprevalence for scrub typhus (61% vs 13%) and a more pronounced age-dependent increase, suggesting more intense and regular exposure. The findings confirm previous studies carried out separately in the two populations and stress the importance of including minority ethnic groups in epidemiological studies on neglected tropical diseases (20, 34). The environmental risk factors associated with seropositivity differed between groups: among Thai participants, exposure to forest or high hill areas and agroforestry fields was associated with higher odds, whereas among highland people, high-risk occupation was the dominant risk factor. In our cohort, neither specific peridomestic risk factors nor preventative behaviours were associated with seropositivity for either population.

In the literature, forests and high hills and, to a lesser extent, agroforestry crops, are well-documented risk environments (17–19, 35). However, despite highland people visiting them more often and reporting worse prevention, these exposures were only significantly associated with seropositivity in the Thai. These findings echo a recent publication from southern India, in which “classical” scrub typhus risk factors were associated with disease only in low-risk populations (11).

Despite no specific agricultural environment being significantly associated with seropositivity in highland people; agricultural workers, day-labourers, and the unemployed or retired (grouped together as high-risk occupations) had higher odds of seropositivity, more so than in the Thai, for which the association was not statistically significant. The high risk among agricultural workers is consistent with the broader literature (18, 35, 36). However, among highland people, day labourers, domestic workers, and unemployed or retired individuals, seroprevalence levels were comparable to agricultural workers. This may reflect exposure to other forms of outdoor activities (e.g. road work, construction) or peridomestic/ everyday life environments that were not explored in our study. Indeed, we found no significant associations with cooking using solid fuels (wood or coal), clearing weeds around the house, or sitting on the lawn or house floor (17, 21, 37). That high-risk occupations had higher odds of seropositivity yet specific environmental associations were not found, suggests a high background risk. Highland people live in areas with densely interspersed agricultural landscapes so individuals must cross many environments in order to reach any specific one; this, coupled with the patchy distribution of scrub typhus transmission foci (38), would obscure specific exposure-outcome relationships. In contrast, the flatlands where most of the Thai population lives have a more homogenous landscape and are therefore less prone to such confounding. In a similar way, small agricultural patches, overgrown vegetation, or abandoned waste may be present near homes and influence risk, yet these factors were not measured in our cohort. Other possible unmeasured confounders include foraging; non-domestic grass harvesting or collection; crop handling, transport and storage; and animal husbandry (16, 19, 36, 39).

We found that exposure to rice fields was associated with lower seroprevalence. This contrasts with most published studies from rural areas, which did not identify strong associations between rice-field exposure and scrub typhus (16, 18, 40). The negative association may reflect non-measured confounders. For example, rice fields are generally well-tended and have less grass overgrowth than other environments. Moreover, rice-field work occurs year-round and encompasses a wide range of activities (e.g. flooding, ploughing, planting, seedling spreading, harvesting), many of which are automated; whereas work in agroforestry plantations is typically manual and concentrated during the rainy and early winter months, when scrub typhus incidence is highest.

We did not find significant associations between seropositivity and preventive behaviours, nor with the overall quality of prevention. This may be due to generally well-implemented prevention, limiting the statistical power, but may also reflect limitations in how behaviours were measured. For instance, we did not ask whether participants wore clothing that was tight on the waist, ankles, and wrists, all of which are entry points for vectors. It is also possible that preventive practice is insufficient to offset the intensity or frequency of exposure in high-risk settings. Indeed, even slight deviations from perfect personal protection have been associated with infection (41). Future efforts should focus not only on promoting prevention but also on deepening our understanding of exposure pathways and raising disease awareness among those who cannot avoid exposure or for whom preventive measures are difficult to implement (22).

### Limitations

In this project, antibodies were measured as a proxy for exposure; but because they can persist above diagnostic cut-off levels for over a year and remain detectable for over ten years, seroconversion could have occurred long before sampling, potentially diluting the measured effect of analysed exposures (42, 43). Moreover, data collection spanned a 9-month period, and because scrub typhus is seasonal, recent exposure will have been more likely in participants sampled during the rainy season. Adding village as a random effect will have reduced seasonality bias.

Because the study questionnaire referred to the preceding 12 month, there was a potential for recall bias. However, agricultural work is fairly constant across years and participants were unaware of their seropositivity status, so the impact should be negligible and non-differential. Prospective cohorts with detailed exposure history could help verify our findings.

Importantly, seroconversion does not equate to clinically relevant disease and should therefore only be used to estimate the impact of risk mitigation campaigns with caution. Indeed, a recent study in high-risk areas of Tamil Nadu (India) indicated that roughly 90% of infections are asymptomatic, much higher than previously thought (2-5%) (44).

Because the primary aim of the HHS project was to estimate the prevalence of several communicable and non-communicable diseases, not risk factors for scrub typhus, the power to detect moderate effects in uncommon exposures (e.g., poor preventive behaviour) was limited, as reflected in the confidence intervals.

Despite efforts to conduct the surveys during periods of low agricultural activity; compared to the official population in the source districts, men (40% in out cohort vs 49% in the population of participating districts) were underrepresented, as were adults aged less than 65 years (75% of all adults vs 84%), this was more marked in the Thai and is probably due to the relatively low compensation offered to participants, which was set at about 30% of a daily labourer’s wage.

Our findings underscore that the strength and nature of scrub typhus risk factors can vary considerably within relatively small geographic areas and between different ethnic groups. Critically, our understanding remains limited among the highland populations, who are at higher risk. Qualitative methods and ethnographic approaches, such as in-depth interviews and participant observation, could be used to improve our understanding, as has been done for other vector-borne diseases (29, 30). Such studies should then inform the development of population-specific messaging and prevention strategies (26). Only through concerted efforts to understand risk, promote awareness and implement context-appropriate prevention can the scrub typhus burden be reduced in these vulnerable communities.

## Conclusions

In rural Chiang Rai, scrub typhus exposure is high, and the burden falls disproportionately on highland people. Because classical exposures poorly predict their risk and their Thai-language proficiency is limited, prevention campaigns delivered in Thai and centred on conventional risk profiles are likely to have limited impact and may perpetuate health inequities(26).

Greater effort is needed to strengthen our understanding of the epidemiology of scrub typhus in high-risk communities and to inform targeted interventions. Qualitative methods, community engagement, and ethnographic approaches are likely to help achieve such goals.

## Ethics statement

The project was approved by the Chiang Rai Public Health Office Ethics Committee (CR-PHO EC, reference number 75/2565) and the Oxford Tropical Research Ethics Committee (OxTREC, reference number 6-22).

Written informed consent was obtained from all participants or their parants/ legal guardians, in addition to the consent of parents/ legal guardians, children provided written informed assent.

## Data availability statement

Data will be made available upon reasonable request to the MORU data management committee.

## Supporting information

**S1 Fig. Venn diagram of agricultural environment exposures**

**S1 Fig. Frequency of preventative practice behaviours among those who visited agricultural environments**

**S1 – Table. Multivariable mixed effects logistic regression model on duration of exposure to agricultural environments and seropositivity**

**S1 Table - Mixed effects logistic regression of seropositivity with village as random intercept**

**S3 Table. Mixed effects logistic regression analysis of preventative behaviours (A), overall prevention quality (B), and domestic/peridomestic risks (C) and seropositivity in highland and Thai participants.**

**S4-Questionnaire** (Scrub typhus questions only)

